# Dry loop–mediated isothermal amplification assay for detection of SARS-CoV-2 from clinical specimens

**DOI:** 10.1101/2020.09.29.20204297

**Authors:** Yuki Higashimoto, Masaru Ihira, Yoshiki Kawamura, Masato Inaba, Kazuya Shirato, Tadaki Suzuki, Hideki Hasegawa, Tsutomu Kageyama, Yohei Doi, Tadayoshi Hata, Tetsushi Yoshikawa

**Affiliations:** Faculty of Medical Technology, Fujita Health University School of Health Sciences, Toyoake, Aichi, Japan; Faculty of Clinical Engineering, Fujita Health University School of Health Sciences, Toyoake, Aichi, Japan; Department of Pediatrics, Fujita Health University School of Medicine, Toyoake, Aichi, Japan; Department of Infectious Diseases, Fujita Health University School of Medicine, Toyoake, Aichi, Japan; Department of Virology III, National Institute of Infectious Diseases, Musashimurayama, Tokyo, Japan; Department of Pathology, National Institute of Infectious Diseases, Musashimurayama,Tokyo, Japan; Influenza Virus Research Center, National Institute of Infectious Diseases, Musashimurayama, Tokyo, Japan; Department of Clinical Laboratory, Fujita Health University Hospital, Toyoake, Japan

**Author notes:** Corresponding Author (YH).

## Abstract

Coronavirus disease 2019 (COVID-19) has had a major disease burden on many countries around the world. The spread of COVID-19 is anticipated to have a major impact on developing countries including African nations. To establish a point-of-care test for COVID-19, we developed a dry loop–mediated isothermal amplification (LAMP) method to detect severe acute respiratory syndrome coronavirus 2 (SARS-CoV-2) RNA. We carried out reverse transcription (RT)-LAMP using the Loopamp SARS-CoV-2 Detection kit (Eiken Chemical, Tokyo, Japan). The entire mixture except for the primers is dried and immobilized inside the tube lid. To determine the specificity of the kit, 22 viral genomes associated with respiratory infections, including the SARS coronavirus, were tested. No LAMP product was detected in reactions performed with RNA from these pathogens. The sensitivity of this assay, determined by either a real-time turbidity assay or colorimetric change of the reaction mixture, as evaluated by the naked eye or under illumination with ultraviolet light, was 10 copies/reaction. After the initial validation analysis, we analyzed 24 nasopharyngeal swab specimens collected from patients suspected to have COVID-19. Nineteen (79.2%) of the 24 samples were positive for SARS-CoV-2 RNA, as determined by real-time RT-PCR analysis. Using the Loopamp SARS-CoV-2 Detection kit, we detected SARS-CoV-2 RNA in 15 (62.5%) of the 24 samples. Thus, the sensitivity, specificity, positive predictive value, and negative predictive value of the Loopamp 2019-CoV-2 detection reagent kit were 94.0%, 96.0%, 95.9%, and 94.1%, respectively. The dry LAMP method for detection of SARS-CoV-2 RNA was fast and easy to use, solves the cold chain problem, and therefore represents a promising tool for diagnosis of COVID-19 in developing countries.

**Author summary:** Coronavirus disease 2019 (COVID-19) is a major public health problem around the world. A reliable point-of-care (POC) test for severe acute respiratory syndrome coronavirus 2 (SARS CoV-2) is urgently needed, especially in developing countries. The loop-mediated isothermal amplification (LAMP) method amplifies template nucleotides under isothermal conditions with high efficiency and specificity, both of which are major advantages for a POC test. In addition, because dry LAMP reagents can be stored at 4°C, it is suitable for use in developing countries.

We evaluated the specificity and sensitivity of the Loopamp SARS-CoV-2 Detection kit (Eiken Chemical, Tokyo, Japan), a dry LAMP method for amplifying viral RNA. The initial validation study revealed that the method was highly specific and sensitive (lower detection limit: 10 copies/reaction). We then analyzed 24 nasopharyngeal swab specimens from patients suspected to have COVID-19. Using the Loopamp SARS-CoV-2 Detection kit, SARS-CoV-2 RNA was detected in 15 (62.5%) of the 24 samples. Compared with the standard real-time reverse transcription PCR, the sensitivity, specificity, positive predictive value, and negative predictive value of the Loopamp SARS-CoV-2 Detection kit were 78.9%, 100%, 100%, and 55.6%, respectively.

## Introduction

The outbreak of coronavirus disease 2019 (COVID-19), caused by the novel coronavirus designated as severe acute respiratory syndrome coronavirus 2 (SARS-CoV-2), started in Wuhan, China, in December 2019 [1, 2]. To date, the outbreak has spread rapidly around the world [1, 2], and WHO has declared it a pandemic. Clinical trials have evaluated several drugs to date, with remdesivir and systemic corticosteroids showing promise for moderate and severe diseases, respectively [3]. Several characteristic clinical findings, such as fever and dry cough with typical signs of pneumonia on chest computed tomography [4–6], history of close contact with a patient or visits to endemic areas [7, 8], are useful in diagnosing COVID-19. However, many patients have mild symptoms or are asymptomatic, hampering accurate diagnosis based on clinical features [9–11]. Therefore, a rapid diagnostic method is necessary in order to provide definitive diagnosis of COVID-19.

Real-time reverse transcription–polymerase chain reaction (RT-PCR) is widely used for diagnosis of COVID-19 [12, 13]. The method is very useful for testing large numbers of samples at large hospitals, diagnostic companies, and local health facilities. However, a point-of-care (POC) test for COVID-19 is also important for management of suspected patients in less resourced settings. In addition, because expansion of COVID-19 to developing countries is a major public health concern, there is an urgent need to develop rapid diagnostic tests for COVID-19. Real-time RT-PCR requires a special thermal cycler with precision optics that monitor fluorescence emission from sample wells. By contrast, the loop-mediated isothermal amplification (LAMP) method can amplify template nucleotides under isothermal conditions with efficiency and specificity as high as those of nested double PCR [14]. Due to its speed, ease of use, and cost-effectiveness, LAMP has been widely used for POC testing for various infectious diseases, including COVID-19 [14–17]. Furthermore, the dry LAMP reagent mixture, which can be stored at 4°C, is much easier to handle and more heat-stable than liquid reagents. Therefore, the dry LAMP method would very useful for diagnosis of tropical infectious diseases in regions including Africa [18], which is expected to be the next epicenter of COVID-19. The aim of this study was to evaluate the performance of the LAMP method using dry reagents for rapid diagnosis of COVID-19 infection. Although several studies have already been published on the use of LAMP to detect SARS-CoV-2 [19–21], this is the first study to evaluate the performance of dry LAMP reagents for this purpose.

## Materials and methods

### Viruses and RNA for initial validation analysis

A total of 22 respiratory pathogens were used for the initial validation analysis of specificity of the primers in this study. Twenty-two viral genomes are listed in Table 1. The Middle East respiratory syndrome coronavirus (MERS-CoV) EMC strain was kindly provided from Dr. Ron A. M. Fouchier, Erasmus Medical Center, Rotterdam, the Netherlands. The SARS-CoV Frankfurt 1 strain was kindly provided from J. Ziebuhr, University of Würzburg, Germany. The clinical isolates of Human coronaviruses (HCoV)-HKU1, OC43, NL63, and 229E were described by previous study [22–24]. *In vitro* transcribed RNA (GenBank accession number MN908947), synthesized using ScriptMax Thermo T7 Transcription Kit (TOYOBO, Osaka, Japan), was used to determine the detection limit of the Loopamp SARS-CoV-2 Detection kit (Eiken Chemical, Tokyo, Japan).

**Table 1.**
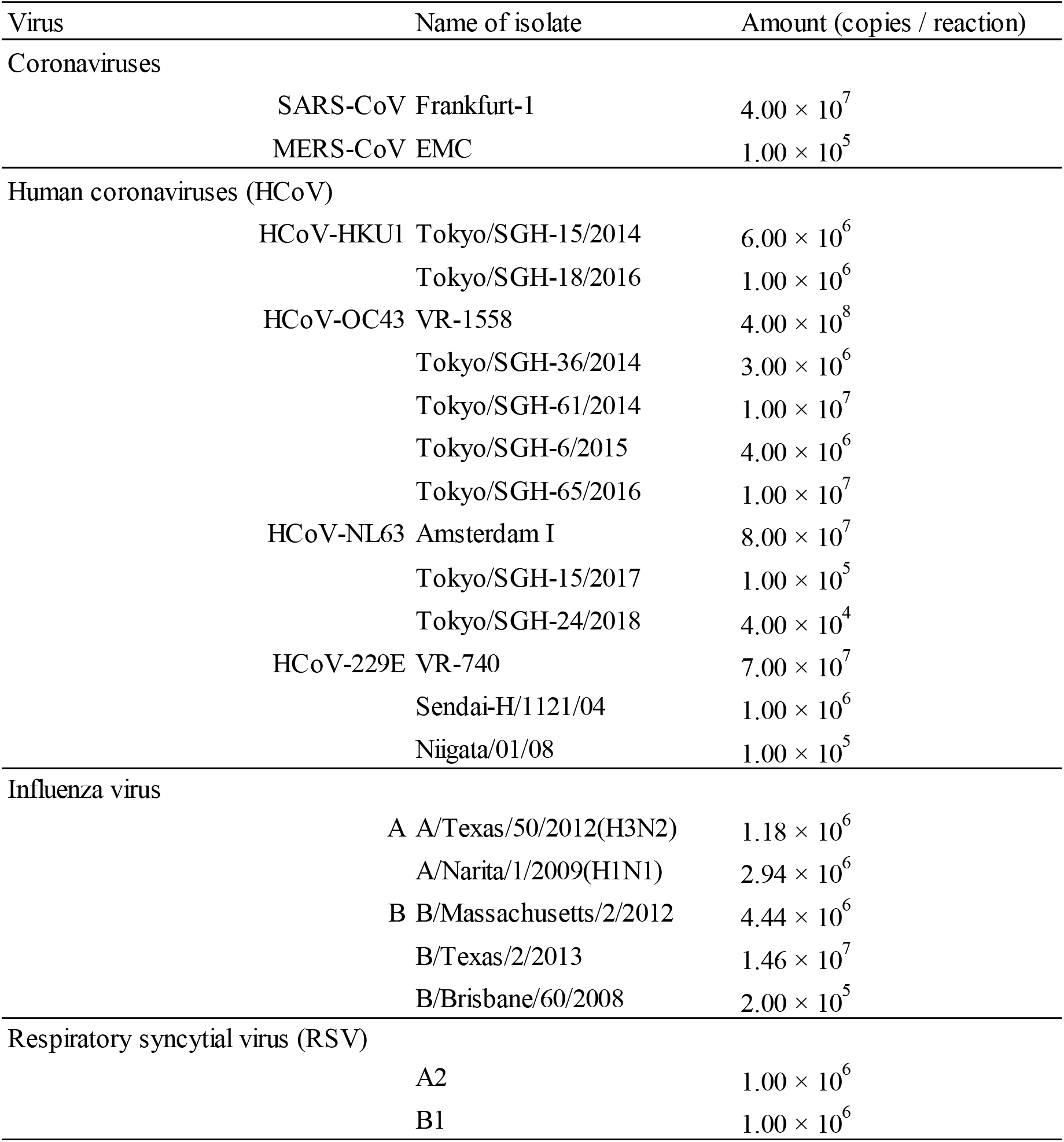
**Specificity of dry loop-mediated isothermal amplification assay for SARS-CoV-2**.

### Clinical specimens

From March 7 to April 30, 2020, nasopharyngeal swabs were collected from patients suspected to have COVID-19 at a university hospital in Japan. Swab samples were collected using a flocked sterile plastic swab applicator and placed in 3 mL of BD universal viral transport medium (Becton, Dickinson and Company, Franklin Lakes, NJ, USA). RNA was extracted from the swab samples immediately. This study was approved by the institutional review board of Fujita Health University (No. HM19-493). Written informed consent was obtained from each patient.

### RNA extraction

Viral RNA was extracted from 140 µL of BD universal viral transport medium in which a nasaopharyngeal swab was immersed. RNA extraction was performed using the QIAamp Viral RNA mini kit (QIAGEN, Chatsworth, CA, USA). After extraction, RNA was eluted in 60 μL of buffer AVE and stored at −80°C.

### Real-time RT-PCR

Real-time RT-PCR assays for detecting SARS-CoV-2 were performed using TaqMan Fast Virus 1-Step Master Mix (Thermo Fisher Scientific, Waltham, MA, USA). Primers and probes were as follows: NIID_2019-nCOV_N_F2, 5’-AAATTTTGGGGACCAGGAAC-3’; NIID_2019-nCOV_N_R2, 5’-TGGCAGCTGTGTAGGTCAAC-3’; NIID_2019-nCOV_N_P2; 5’-FAM ATGTCGCGCATTGGCATGGA BHQ-3’ [25]. Single-well denaturation, reverse transcription, and amplification steps were performed on a QuantStudio 1 Real-Time PCR System (Thermo Fisher Scientific, Waltham, MA, USA) in the standard mode. Primer and probe concentrations were as follows: NIID_2019-nCOV_N_F2, 500 nM; NIID_2019-nCOV_N_R2, 700 nM; NIID_2019-nCOV_N_P2, 200 nM. PCR conditions were as follows: RT at 50 °C for 5 min; enzyme activation at 95 °C for 20 sec; and 45 cycles of denaturation at 95 °C for 15 sec and primer annealing/extension/fluorescence emission at 60°C for 60 sec. The real-time RT-PCR reaction mixture (20 µL total volume) contained 5.0 µL of 4× Fast Virus Master Mix, 1.0 µL of primer–probe pre-mix, 5.0 µL of template RNA and nuclease-free water.

### Reverse Transcription–LAMP

Reverse transcription (RT)-LAMP was carried out using the Loopamp SARS-CoV-2 Detection kit (Eiken Chemical). Because the entire mixture except the primers is dried and immobilized inside the tube lid, 10 microliters of purified RNA and 15 microliters of SARS-CoV-2 specific primer sets were added to the bottom of the tube, and then the tube was inverted several times to resuspend the enzyme and buffer. The full reaction mixture was collected at the bottom of the tube by a quick spin-down. The mixture was incubated in a real-time turbidimeter (LA-200; Eiken Chemical) for 35 min at 62.5°C. For visual evaluation of fluorescence, the reaction tube was illuminated with ultraviolet light using an ultraviolet illumination system (WSE-5300; ATTO, Tokyo, Japan) and and also observed by the naked eye.

## Results

### Specificity and sensitivity of LAMP methods

To determine the specificity of the Loopamp SARS-CoV-2 Detection kit, we tested 22 viral genomes including SARS coronavirus, Middle East Respiratory Syndrome (MERS) coronavirus, other human coronaviruses, influenza viruses, and respiratory syncytial viruses associated with respiratory infections (Table 1). No LAMP product was detected in reactions performed with RNA from these pathogens. These results were confirmed by a turbidity assay and agarose gel electrophoresis analysis (data not shown). To determine the sensitivity of the Loopamp SARS-CoV-2 Detection kit, *in vitro* transcribed RNAs were serially diluted in 10 mM Tris buffer containing 0.1 mM EDTA and 50 ng/mL of carrier RNA were used to define the detection limit. The sensitivity of this assay, determined by either the turbidity assay or colorimetric change of the reaction mixtures evaluated by the naked eye, was 10 copies/reaction (Fig. 1).

**Figure 1.**
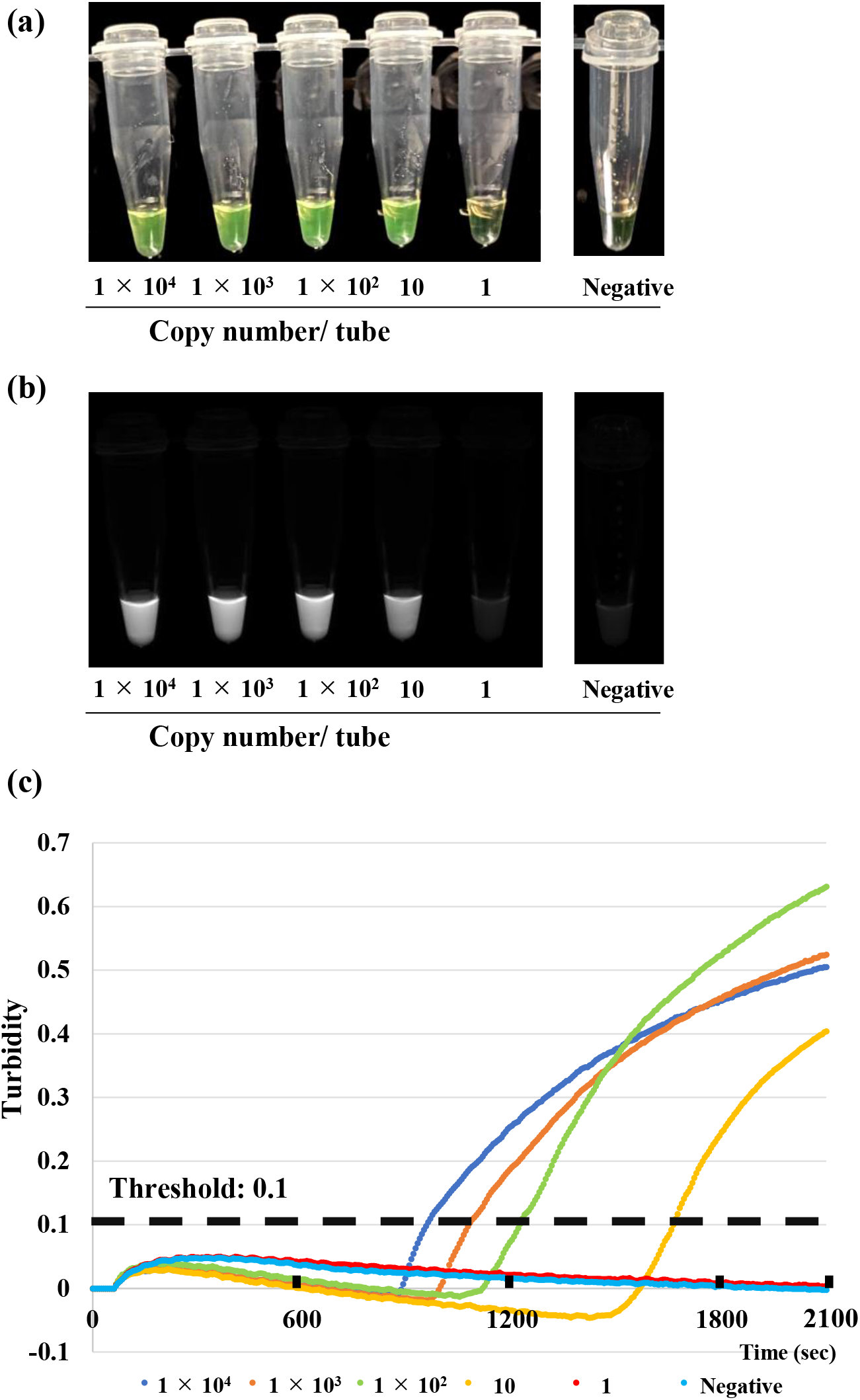
Sensitivity of the SARS-CoV-2 dry LAMP assay, determined using serially diluted *in vitro* transcribed RNA. (a) Naked-eye detection at the end of assay (35 min). Green indicates a positive reaction, and orange indicates a negative reaction. (b) Ultraviolet light detection at the end of the assay (35 min). Light gray indicates a positive reaction, and dark gray indicates a negative reaction. (C) Real-time turbidity assay.

### Evaluation of clinical applications

To evaluate the performance of the Loopamp SARS-CoV-2 Detection kit as a POC test, we analyzed 24 nasopharyngeal specimens collected from patients suspected of having COVID-19, including three asymptomatic individuals who came into close contact with COVID-19 patients (Table 2). Nineteen (79.2%) of the 24 samples were positive for SARS-CoV-2 RNA by real-time RT-PCR analysis. Using the Loopamp SARS-CoV-2 Detection kit, SARS-CoV-2 RNA was detected in 15 (62.5%) of the 24 samples. The four false-negative samples contained low copies of viral RNA (2.6, 2.3, 2.2, and 0.5 copies/reaction) and were collected during the convalescent phase of illness (days 7 to 20). Thus, the sensitivity, specificity, positive predictive value, and negative predictive value of the Loopamp SARS-CoV-2 Detection kit were 78.9%, 100%, 100%, and 55.6%, respectively.

**Table 2.** **Comparison of real-time RT-PCR and dry loop–mediated isothermal amplification assay results of SARS-CoV-2 using clinical specimens**.

| Case | Sampling day* | Real-time RT-PCR<br>(copies/ reaction) | LAMP |
| --- | --- | --- | --- |
| 1 | unknown** | 212732.5 | + |
| 2 | unknown** | 94749.3 | + |
| 3 | 7 | 22910.9 | + |
| 4 | 5 | 3796.9 | + |
| 5 | 11 | 1544.6 | + |
| 6 | 10 | 946.9 | + |
| 7 | 8 | 413.9 | + |
| 8 | 8 | 280.8 | + |
| 9 | 9 | 146.0 | + |
| 10 | unknown** | 68.7 | + |
| 11 | 12 | 49.8 | + |
| 12 | 12 | 48.9 | + |
| 13 | 7 | 14.2 | + |
| 14 | 8 | 13.2 | + |
| 15 | 19 | 4.4 | + |
| 16 | 7 | 2.6 | - |
| 17 | 18 | 2.3 | - |
| 18 | 10 | 2.2 | - |
| 19 | 20 | 0.5 | - |
| 20 | 14 | 0.0 | - |
| 21 | 8 | 0.0 | - |
| 22 | 12 | 0.0 | - |
| 23 | 10 | 0.0 | - |
| 24 | 12 | 0.0 | - |

\* Onset of disease was defined as day 1.
\*\* Sampling day was unknown because the patient was asymptomatic.

## Discussion

Ease of use, speed, and low cost of amplifying target nucleotides is a significant advantage of the LAMP assay as a POC test. Accordingly, several investigators have developed SARS-CoV-2 LAMP assays [16, 19–21, 26]. To further leverage the advantages of the LAMP method, several improvements have been reported, including colorimetric detection using the naked eye [20] or fluorescent detector [26] and direct detection of target sequence without RNA extraction. These improvements shorten and simplify the workflow of the LAMP method [20], and enable high-throughput analysis. Using the LAMP method with dry reagents allows the use of this assay in developing countries, as the reagents can be stored in refrigerators that overcomes the requirement for strict cold-chain transportation and storage of the reagents. Therefore, in this study, we investigated the performance of the SARS-CoV-2 dry LAMP method.

No amplification was observed with other viral genomes (including SARS coronavirus, MARS coronavirus, and other human coronaviruses associated with respiratory infections) using the SARS-CoV-2 dry LAMP method (Table 1), indicating that this LAMP assay can specifically amplify SARS-CoV-2. In addition, the detection limit of the kit, based on the turbidity assay and colorimetric change determined by the naked eye, was 10 copies/reaction, almost the same or slightly higher than the previously reported SARS-CoV-2 LAMP assays [26]. These initial validation analysis demonstrated that the SARS-CoV-2 dry LAMP method is highly specific and sensitive. Huang et al. have shown that SARS-CoV-2 RNA can be amplified by the LAMP assay without RNA extraction [20], in line with our previous studies for detection of herpes simplex virus [27] and human herpesvirus-6 [28]. Therefore, to further improve the LAMP method as a POC test, a direct detection method should be developed.

In order to evaluate the performance of SARS-CoV-2 dry LAMP for analyzing clinical samples, we analyzed 24 nasopharyngeal swab specimens collected from patients suspected of having COVID-19. The sensitivity and specificity of this assay was sufficient for practical purposes. In 24 nasopharyngeal swab specimens, there are only 5 SARS-CoV-2 RNA negative samples. While there were 4 false-negative test results, as shown in Table 2, they represented specimens with the lowest copy numbers. In particular, two of the four false-negative samples were collected on days 18 and 20 to confirm the absence of viral genome prior to hospital discharge. Given that all samples with more than 4.4 copies/reaction were detected by both turbidity assay and colorimetric changes, we believe that this SARS-CoV-2 dry LAMP method is reliable for clinical use in diagnosing COVID-19.

In summary, this study demonstrated that the SARS-CoV-2 dry LAMP method is a reliable method for rapid diagnosis of COVID-19. The dry LAMP method overcomes the requirement for strict cold-chain transportation and storage of reagents. Therefore, this method is expected to be a useful POC test in developing countries where COVID-19 is spreading.

## Data Availability

All data generated or analysed during this study are included in this published article.

## Acknowledgments

We gratefully acknowledge Eiken Chemical for their contributions to this work.

